# Development and Validation of a Nomogram for Predicting False Negative IGRA Results in Pulmonary Tuberculosis Patients Using Propensity Score Matching

**DOI:** 10.1101/2025.01.05.25320016

**Authors:** Feng Zhang, Yong Gao, Tuantuan Li, Wei Zhang

## Abstract

**Objective:** Exploration of Factors Influencing False-Negative Results in Interferon-Gamma Release Assay (IGRA) for Patients with Pulmonary Tuberculosis (PTB), and Development of a Nomogram Model to Predict IGRA False Negatives, to Optimize Clinical Diagnosis and Treatment Decisions.

**Methods:** A total of 143 patients diagnosed with Pulmonary Tuberculosis (PTB) were selected for this study. Among them, 63 patients who were IGRA negative but positive for pathogen detection formed the observation group, while 80 patients who were both IGRA positive and pathogen positive constituted the control group. After balancing potential confounding factors between the two groups using Propensity Score Matching (PSM), clinical characteristics and laboratory indicators of the two groups were compared. Logistic regression analysis was then employed to identify independent risk factors affecting IGRA results. Based on significantly associated factors, a nomogram model was constructed, and its predictive performance was evaluated.

**Results:** After propensity score matching, each group consisted of 55 patients. Compared to the control group, the observation group showed significant differences in white blood cell count (WBC), neutrophil count (NEUT), lymphocyte count (LYM), red blood cell count (RBC), hemoglobin (HGB), and albumin (ALB) levels (*P* < 0.05). Logistic regression analysis revealed that RBC and ALB were influencing factors for false-negative IGRA results. The constructed nomogram model demonstrated a good fit (χ^2^=6.444, *P*=0.598), with an area under the receiver operating characteristic curve (AUC) of 0.703 (95% CI: 0.605-0.800), accuracy of 0.682 (95% CI: 0.586-0.767), sensitivity of 0.691 (95% CI: 0.569-0.813), specificity of 0.673 (95% CI: 0.549-0.797), positive predictive value (PPV) of 0.679 (95% CI: 0.556-0.801), and negative predictive value (NPV) of 0.685 (95% CI: 0.561-0.809). Decision curve analysis indicated that the net benefit of predicting false-negative IGRA results using this nomogram model was greater than 0 when the threshold probability ranged from 0.15 to 0.75.

**Conclusion:** Lower levels of RBC and ALB may be significant factors contributing to false-negative IGRA results in PTB patients. The constructed nomogram model, incorporating these factors, holds considerable clinical application value for predicting IGRA false negatives, aiding in the improvement of early diagnosis and management strategies for PTB.

## Introduction

Pulmonary Tuberculosis (PTB), caused by *Mycobacterium tuberculosis*, is a chronic infectious disease that poses a significant threat to global public health and remains one of the leading causes of death from infectious diseases worldwide [1,2].According to the latest report from the World Health Organization (WHO), millions of people worldwide are infected with and die from PTB each year, with particularly high incidence and mortality rates in developing countries. Notably, China is among the high-burden countries for PTB, ranking third globally in terms of incidence. In China, the estimated number of PTB cases is 748,000, resulting in an incidence rate of 52 per 100,000 population [3].Early diagnosis plays a vital role in controlling the transmission of PTB and is essential for improving patient outcomes. Timely detection not only aids in the prompt initiation of appropriate treatment but also helps prevent the further spread of the disease within communities [4].Interferon-Gamma Release Assays (IGRAs) represent a novel in vitro immunological testing method that has been widely adopted for the auxiliary diagnosis of PTB. This is due to their high specificity and the advantage of not being affected by Bacille Calmette-Guérin (BCG) vaccination [5,6].However, despite the many advantages of IGRAs, they are not 100% accurate in clinical settings. Some patients diagnosed with PTB)may exhibit false-negative IGRA results [7-9].

This not only increases the complexity of clinical diagnosis but can also lead to delayed treatment or misdiagnosis, thereby affecting timely patient care and disease control strategies.Given the potential significant impact of false-negative IGRA results on clinical decision-making, this study aims to thoroughly investigate the underlying factors contributing to false-negative IGRA outcomes in PTB patients and to establish a predictive model for identifying such cases. To reduce the influence of confounding factors and ensure robust comparisons, Propensity Score Matching (PSM) was utilized to match groups based on key covariates. Through comprehensive analysis of clinical features and laboratory parameters, this research endeavors to offer clinicians a more accurate diagnostic tool, enhancing early detection and guiding optimized management strategies for PTB patients.

## Materials and Methods

### Study Subjects

This study adopted a retrospective cohort design, recruiting a total of 143 patients diagnosed with PTB at the Second People’s Hospital of Fuyang City, Anhui Province, from January 2023 to September 2024. Patients were categorized into two groups based on their IGRA test results: the observation group comprised 63 patients who tested IGRA negative but were pathogen positive, whereas the control group included 80 patients who tested both IGRA positive and pathogen positive. This approach allowed for a detailed comparison between patients with discordant and concordant IGRA and pathogen test results.The study protocol received approval from the Ethics Committee of the Second People’s Hospital of Fuyang City and complied with the principles outlined in the Helsinki Declaration (Approval No.: 20231112033). Considering the retrospective nature of this research, the need for informed consent was waived, as approved by the Ethics Committee. To safeguard patient privacy, all data were managed in strict compliance with ethical guidelines, ensuring both confidentiality and anonymity. In the analysis and reporting of the study results, no personal identifying information was utilized, thereby protecting the identities of all participants.

### Inclusion and exclusion criteria

Inclusion Criteria: (1)Patients diagnosed with PTB according to the “WS288—2017 Pulmonary Tuberculosis Diagnosis” guidelines [10]; (2)All included patients had a confirmed positive pathogen status verified by the Xper MTB/RIF assay and simultaneously underwent IGRA testing; (3)Patients with complete clinical records and laboratory test data.Exclusion Criteria: (1)Patients with coexisting severe infections, autoimmune diseases, malignancies, organ transplants, or HIV positivity that could affect immune function; (2)Patients who had received immunosuppressive therapy or used steroid medications.

### Patients data collection

The clinical data collected for hospitalized patients included demographic information such as gender, age, and body mass index (BMI), as well as a range of symptoms including cough, sputum production, hemoptysis, dyspnea, loss of appetite, and fever. Medical history details were also gathered, covering smoking and alcohol consumption history, the presence of diabetes mellitus and hypertension, chronic lung diseases such as chronic obstructive pulmonary disease, emphysema, and bronchiectasis, and the presence of pulmonary cavitation.BMI=weight (in kilograms) / (height (in meters))^2.

### Clinical laboratory data

For the blood tests, 5 mL of whole blood was collected in BD vacuum tubes with lithium heparin anticoagulant. One milliliter of blood was added to each of the N, P, and T reaction tubes, which were then incubated at a constant temperature of 37°C for 24 hours. Following incubation, IGRA testing was performed using the Wantai CARIS200 chemiluminescent immunoassay analyzer and its corresponding reagents.2 mL of blood containing EDTA-K2 anticoagulant was drawn and analyzed on a SYSMEX XE2100 automated hematology analyzer along with its corresponding reagent kit to measure white blood cell count (WBC), neutrophil count (NEUT), lymphocyte count (LYM), red blood cell count (RBC), hemoglobin (HGB), and platelet count (PLT).3 mL of fasting morning blood was collected from patients and analyzed on a Hitachi 7600 automated biochemical analyzer to determine albumin (ALB) and C-reactive protein (CRP) levels, The assay kits are from Zhongsheng North Control Biotechnology Co.Based on the collected data, ratios including the Neutrophil-to-Lymphocyte Ratio (NLR) and Platelet-to-Lymphocyte Ratio (PLR) were computed.

For the Xpert MTB/RIF test, 1 to 2 mL of the specimen was transferred to a 15 mL centrifuge tube. Depending on the nature of the specimen, 1 to 2 times the volume of Sample Reagent (SR) was added. The mixture was vortexed for 30 seconds until no clumps remained, then left to stand for 20 minutes, during which it was shaken at least twice. After this period, 2 mL of the thoroughly liquefied specimen was aspirated and transferred to the Xpert MTB/RIF cartridge for analysis [11].

### Statistical Analysis

Statistical analyses were performed using SPSS version 26.0. For continuous data, the Kolmogorov-Smirnov (K-S) test was used to assess normality. Data conforming to a normal distribution were analyzed using independent samples t-tests and expressed as mean ± standard deviation (SD). Data with skewed distributions were analyzed using the Mann-Whitney U test and presented as median (M) with interquartile range [P25, P75]. Categorical data were summarized as counts (n) and percentages (%) and analyzed using the chi-square (χ^2^) test.Propensity score matching was conducted using a 1:1 nearest neighbor method with a caliper value of 0.02 to balance potential confounding factors between the two groups. The dependent variable for matching was whether the IGRA result was false negative, while the covariates included gender, age, BMI, cough, sputum production, hemoptysis, dyspnea, loss of appetite, and fever. Logistic regression analysis was employed to identify independent risk factors for false-negative IGRA results.The nomogram prediction model was constructed using R version 4.4.2 with the “rms” package. Calibration curves, receiver operating characteristic (ROC) curves, and Hosmer-Lemeshow (HL) goodness-of-fit tests were utilized to evaluate the accuracy of the nomogram model using the “pROC” and “rms” packages. Decision curve analysis (DCA) was conducted using the “rmda” package to assess the clinical utility of the prediction model. A p-value less than 0.05 (*P* < 0.05) was considered to indicate statistical significance.

## Results

### Comparison of baseline characteristics before and after matching between the two groups

Before propensity score matching, a statistically significant difference in age was observed between the two groups (*P*<0.05). After successfully matching 55 pairs of patients using PSM, no statistically significant differences were found between the groups for any of the factors (*P*>0.05) (Table 1). The propensity score distribution plot illustrates that the distribution was imbalanced between the two groups before matching but became markedly more consistent after matching, with differences notably reduced (Figure 1).

**Table 1.** Comparison of baseline characteristics before and after propensity matching between the two groups.

| Characteristic | Before propensity score matching |  |  | After propensity score matching |  |  |
| --- | --- | --- | --- | --- | --- | --- |
|  | Observation group (n=80) | Control group (n=63) | *P* value | Observation group (n=55) | Control group (n=55) | *P* value |
| Age[M(P25, P75)] | 71.00(59.00,79.00) | 62.00(51.00,76.00) | 0.028 | 69.00(57.00,78.00) | 69.00(55.00,78.00) | 0.986 |
| Gender[n(%)] |  |  | 0.653 |  |  | 0.621 |
| Male | 53(84.13) | 65(81.25) |  | 46(83.64) | 44(80.00) |  |
| Female | 10(15.87) | 15(18.75) |  | 9(16.36) | 11(20.00) |  |
| BMI(mean±SD) | 19.80±3.10 | 20.44±3.02 | 0.21<br>6 | 20.01±3.24 | 20.38±3.11 | 0.53<br>4 |
| Coughing[n(%)] | 56(88.89) | 69(86.25) | 0.63<br>7 | 49(89.09) | 48(87.27) | 0.76<br>8 |
| Sputum<br>production[n(%)] | 55(87.30) | 65(81.25) | 0.32<br>8 | 48(87.27) | 48(87.27) | 1.00<br>0 |
| Hemoptysis[n(%)] | 5(7.94) | 8(10.00) | 0.67<br>0 | 4(7.27) | 2(3.64) | 0.40<br>1 |
| Dyspnea[n(%)] | 30(47.62) | 28(35.00) | 0.12<br>7 | 24(43.64) | 26(47.27) | 0.70<br>2 |
| Loss of<br>appetite[n(%)] | 26(41.27) | 22(27.50) | 0.08<br>3 | 18(32.73) | 18(32.73) | 1.00<br>0 |
| Fever[n(%)] | 30(47.62) | 35(43.75) | 0.64<br>5 | 27(49.09) | 25(45.45) | 0.70<br>2 |

**Figure 1.**
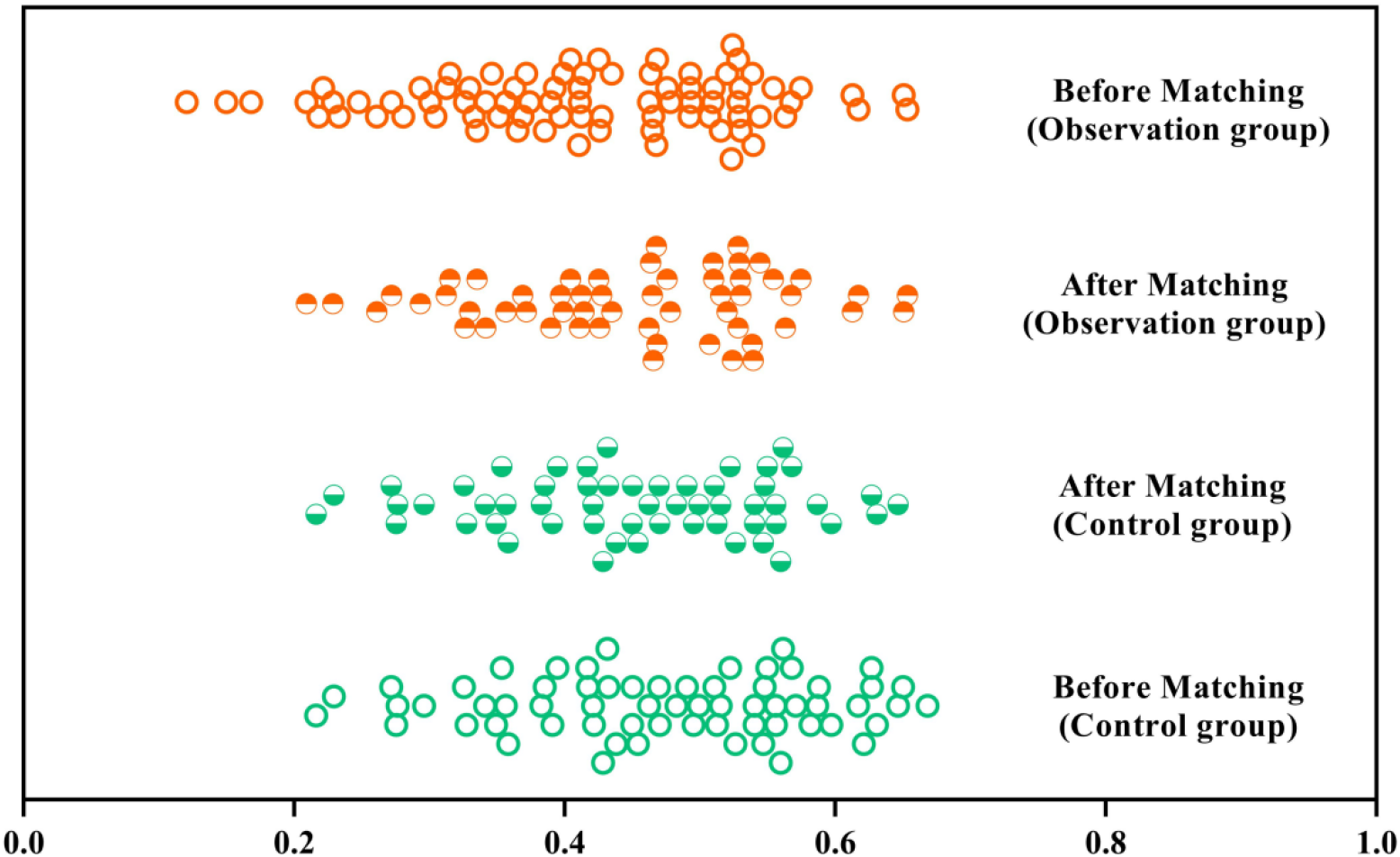
Distribution of scores before and after matching in the two groups

### Comparative analysis of related factors between the two groups after matching

After matching, a comparative analysis was conducted on smoking history, alcohol consumption history, diabetes mellitus, hypertension, chronic lung diseases, pulmonary cavitation, and laboratory indicators between the two groups. The results showed significant differences in WBC, NEUT, LYM, RBC, ALB, and NLR (*P*<0.05). No statistically significant differences were found in the other factors (*P*>0.05) (Table 2).

**Table 2.**
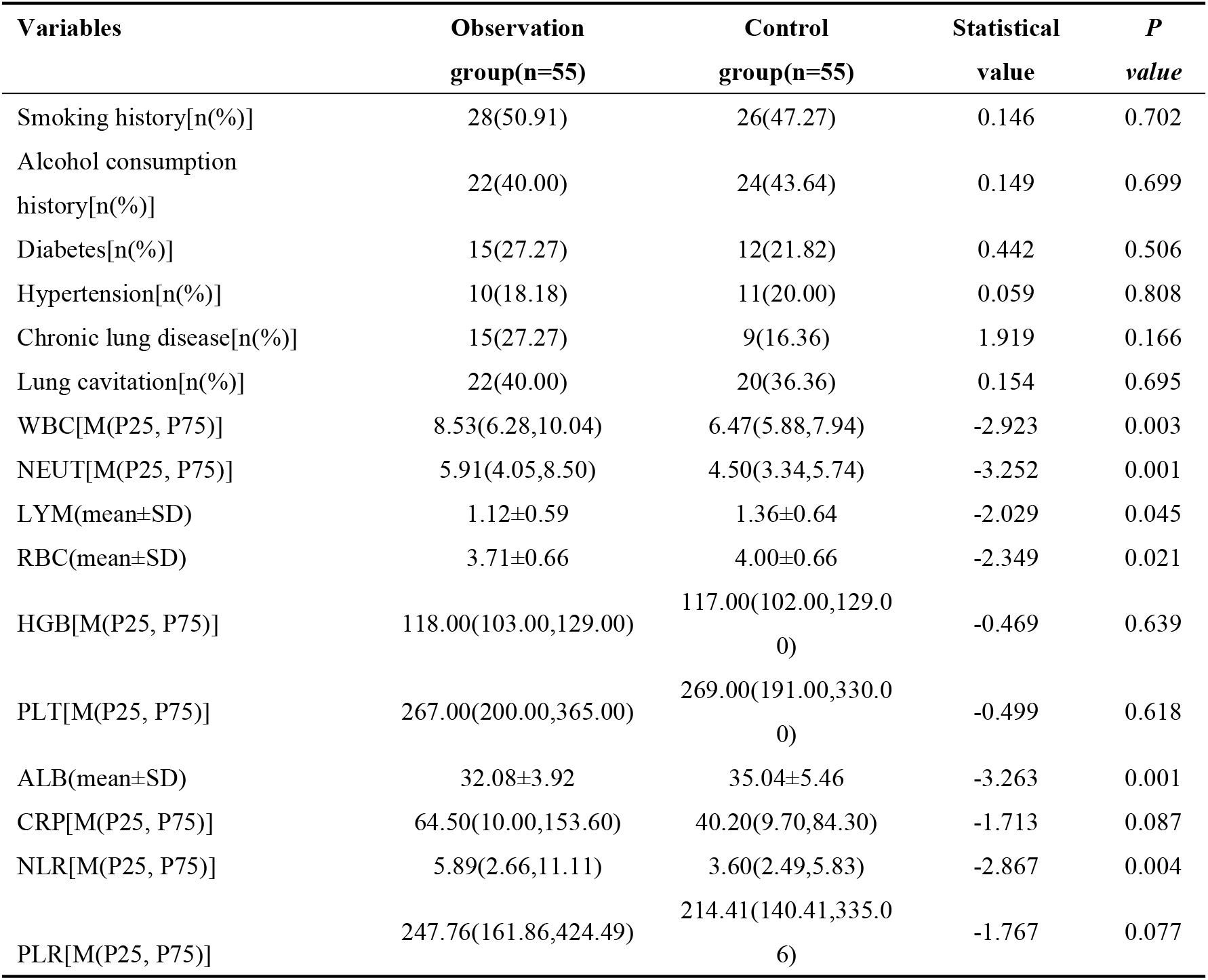
Comparative analysis of related factors between the two groups after matching.

| Variables | Observation<br>group(n=55) | Control<br>group(n=55) | Statistical<br>value | P<br>value |
| --- | --- | --- | --- | --- |
| Smoking history[n(%)] | 28(50.91) | 26(47.27) | 0.146 | 0.702 |
| Alcohol consumption<br>history[n(%)] | 22(40.00) | 24(43.64) | 0.149 | 0.699 |
| Diabetes[n(%)] | 15(27.27) | 12(21.82) | 0.442 | 0.506 |
| Hypertension[n(%)] | 10(18.18) | 11(20.00) | 0.059 | 0.808 |
| Chronic lung disease[n(%)] | 15(27.27) | 9(16.36) | 1.919 | 0.166 |
| Lung cavitation[n(%)] | 22(40.00) | 20(36.36) | 0.154 | 0.695 |
| WBC[M(P25, P75)] | 8.53(6.28,10.04) | 6.47(5.88,7.94) | -2.923 | 0.003 |
| NEUT[M(P25, P75)] | 5.91(4.05,8.50) | 4.50(3.34,5.74) | -3.252 | 0.001 |
| LYM(mean±SD) | 1.12±0.59 | 1.36±0.64 | -2.029 | 0.045 |
| RBC(mean±SD) | 3.71±0.66 | 4.00±0.66 | -2.349 | 0.021 |
| HGB[M(P25, P75)] | 118.00(103.00,129.00) | 117.00(102.00,129.00) | -0.469 | 0.639 |
| PLT[M(P25, P75)] | 267.00(200.00,365.00) | 269.00(191.00,330.00) | -0.499 | 0.618 |
| ALB(mean±SD) | 32.08±3.92 | 35.04±5.46 | -3.263 | 0.001 |
| CRP[M(P25, P75)] | 64.50(10.00,153.60) | 40.20(9.70,84.30) | -1.713 | 0.087 |
| NLR[M(P25, P75)] | 5.89(2.66,11.11) | 3.60(2.49,5.83) | -2.867 | 0.004 |
| PLR[M(P25, P75)] | 247.76(161.86,424.49) | 214.41(140.41,335.06) | -1.767 | 0.077 |

### Multivariate Binary Logistic Regression Analysis

A multivariate binary logistic regression analysis was conducted with IGRA false negativity as the dependent variable and WBC, NEUT, LYM, RBC, ALB, and NLR as independent covariates. The results indicated that RBC and ALB were independent influencing factors for IGRA false negativity (*P*<0.05) (Table 3).

**Table 3.** Multivariate Binary Logistic Regression Analysis.

| Variable | $\beta$ | SE | Wald $\chi^2$ | P value | OR | 95%CI | |
| --- | --- | --- | --- | --- | --- | --- | --- |
|  |  |  |  |  |  | lower limit | upper limit |
| WBC | 0.102 | 0.350 | 0.085 | 0.771 | 1.107 | 0.558 | 2.198 |
| NEUT | -0.094 | 0.421 | 0.050 | 0.823 | 0.910 | 0.399 | 2.075 |
| LYM | 0.365 | 0.640 | 0.325 | 0.568 | 1.441 | 0.411 | 5.051 |
| RBC | -0.892 | 0.393 | 5.160 | 0.023 | 0.410 | 0.190 | 0.885 |
| ALB | -0.127 | 0.047 | 7.101 | 0.008 | 0.881 | 0.803 | 0.967 |
| NLR | 0.199 | 0.112 | 3.181 | 0.075 | 1.220 | 0.980 | 1.518 |

### Development and Validation of the Nomogram Model

A nomogram model was constructed using RBC and ALB as predictors to estimate the risk of IGRA false negativity in pulmonary tuberculosis (PTB) patients (Figure 2). The Hosmer-Lemeshow goodness-of-fit test indicated that the nomogram model had a good fit (*χ*^*2*^=6.444, *P*=0.598) (Figure 3).ROC curve analysis showed an AUC of 0.703 (95% CI: 0.605-0.800), with overall accuracy of 0.682 (95% CI: 0.586-0.767), sensitivity of 0.691 (95% CI: 0.569-0.813), specificity of 0.673 (95% CI: 0.549-0.797), positive predictive value (PPV) of 0.679 (95% CI: 0.556-0.801), and negative predictive value (NPV) of 0.685 (95% CI: 0.561-0.809) (Figure 4).Decision curve analysis demonstrated that the net benefit of the nomogram model for predicting IGRA false negativity remained greater than zero when the threshold probability ranged from 0.15 to 0.75 (Figure 5).

**Figure 2.**
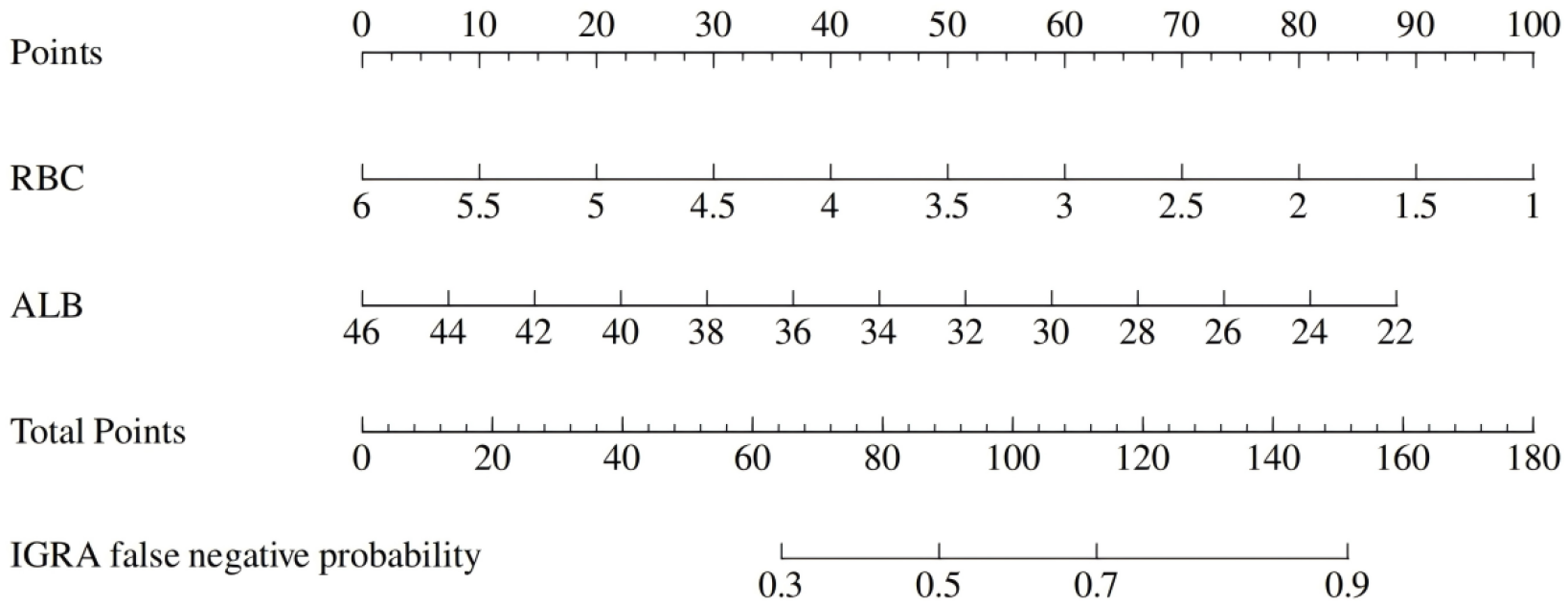
Nomogram model

**Figure 3.**
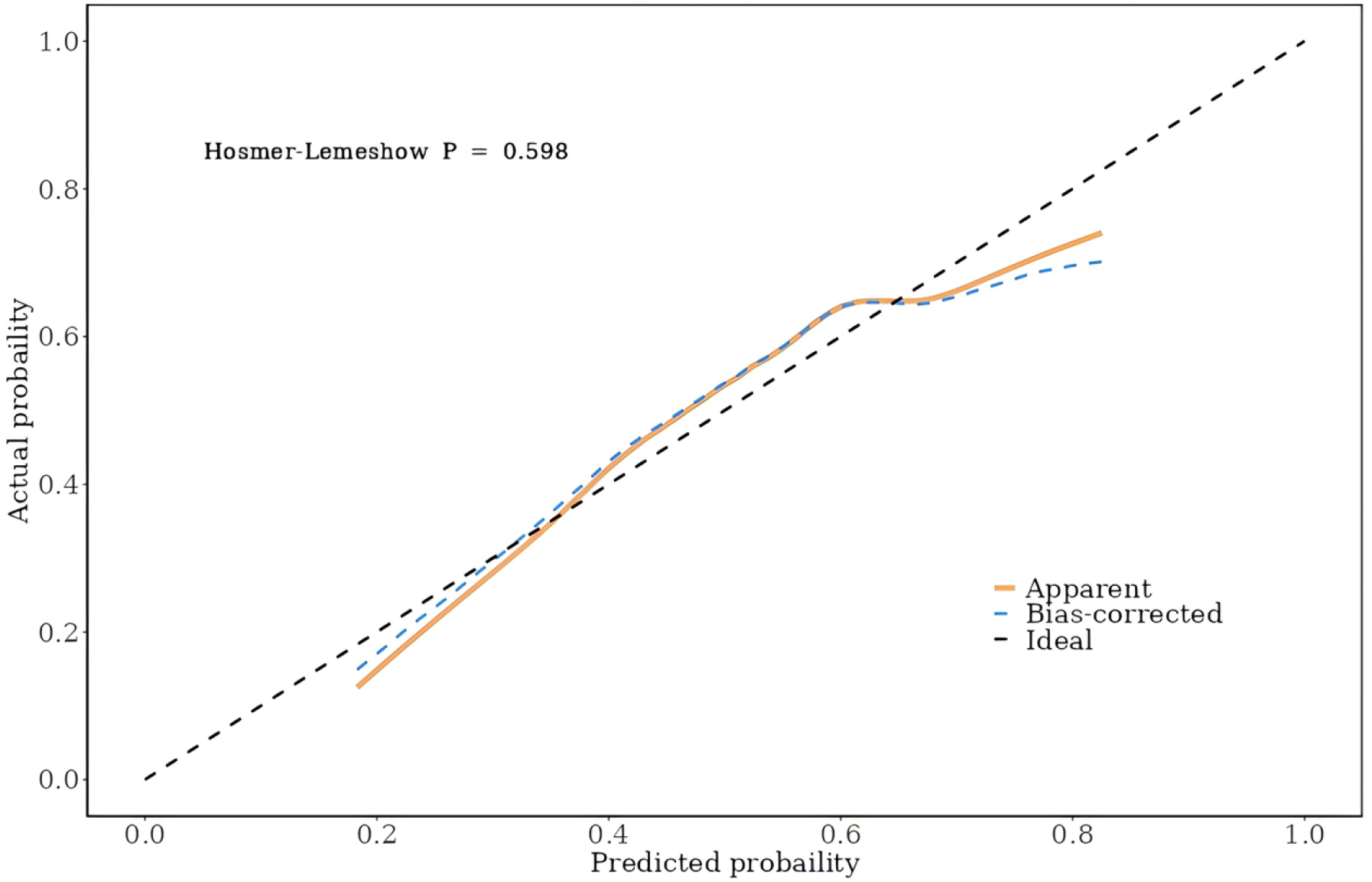
Calibration curve

**Figure 4.**
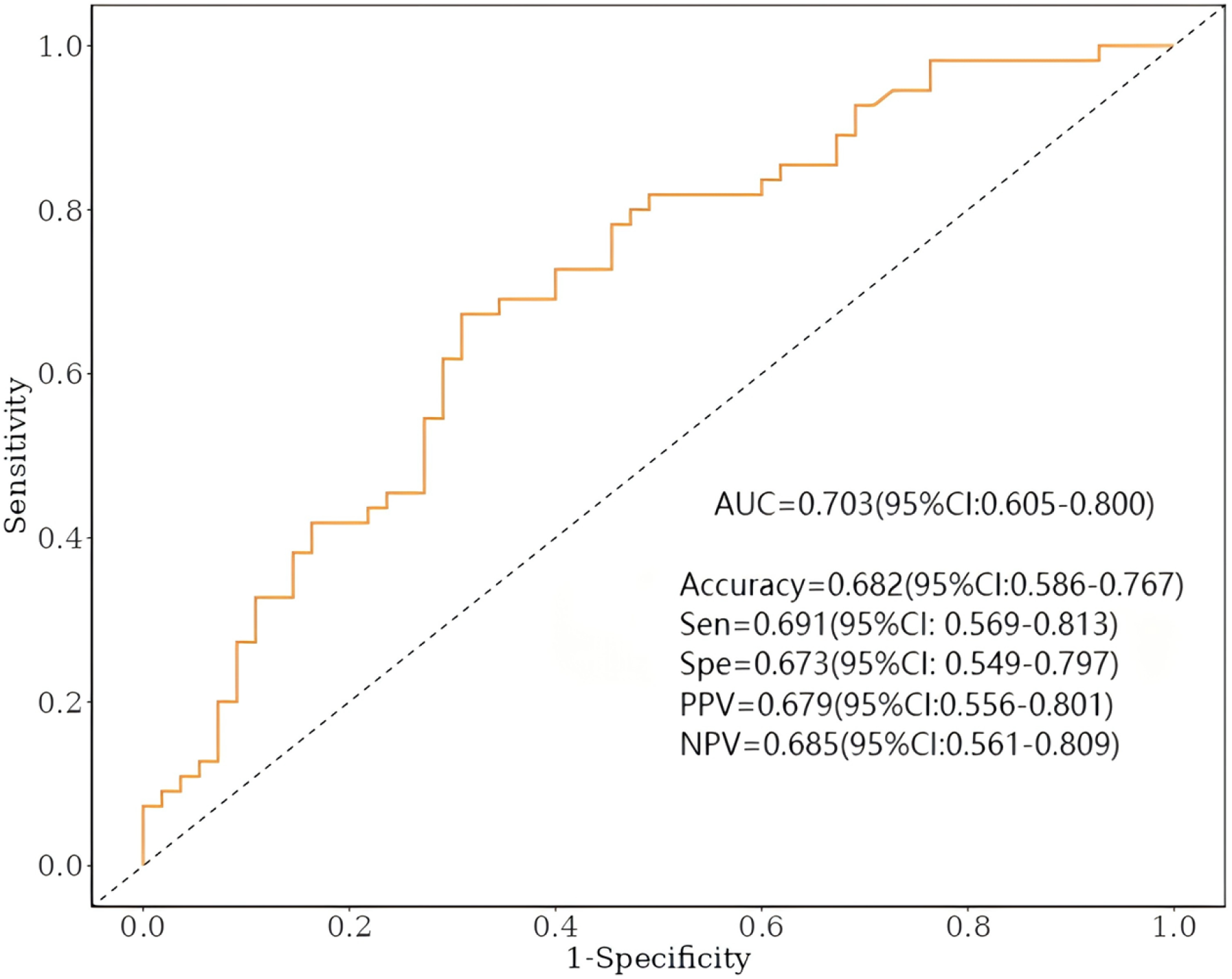
ROC curve

**Figure 5.**
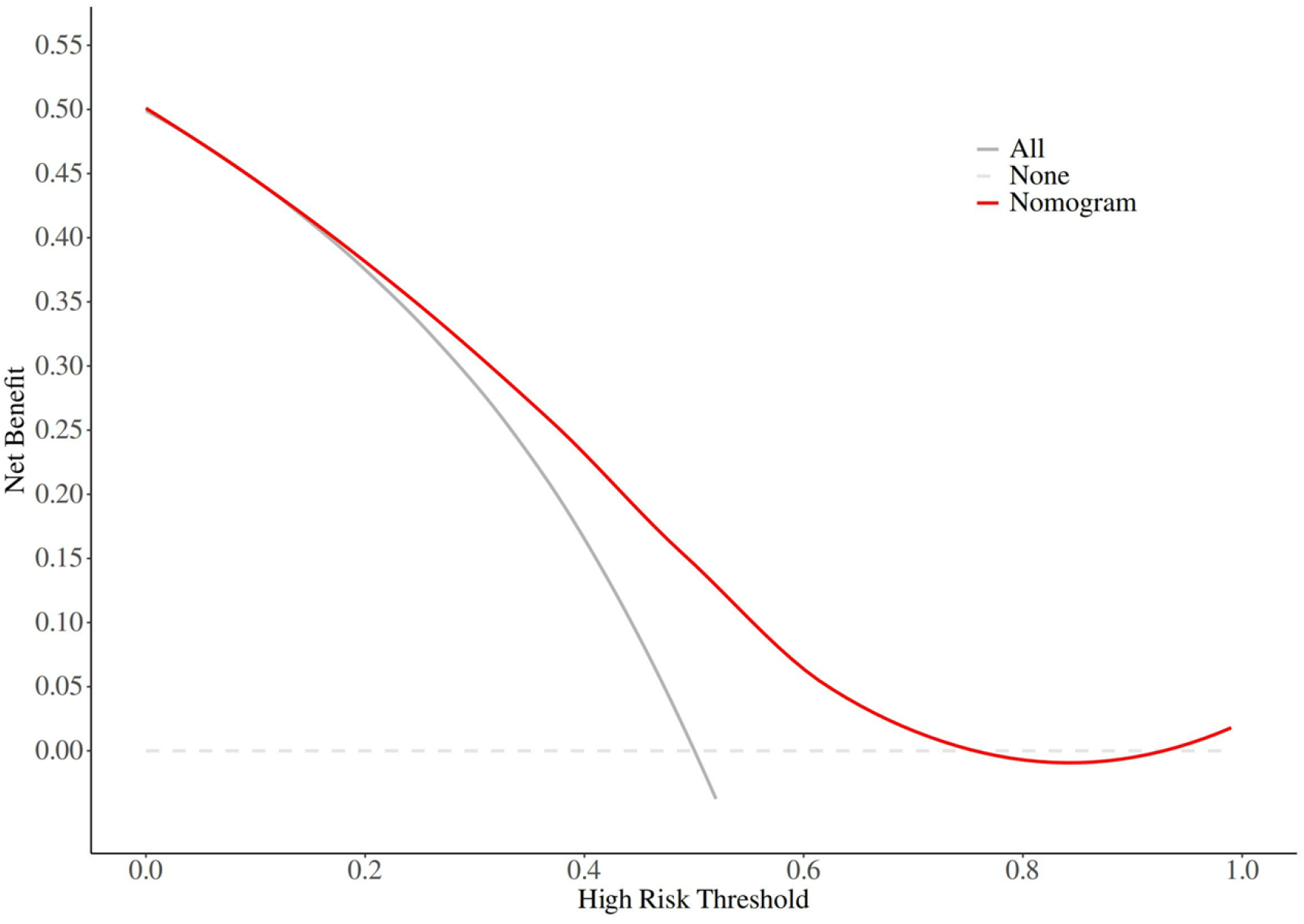
Decision curves

## Discussion

To mitigate the influence of confounding factors on the study results, this research utilized PSM. Traditionally, controlling for confounding variables has often depended on multivariate regression analysis. This method assumes that all potential confounders have been identified and incorporated into the model and requires that the relationships among these factors are accurately specified [12]. However, this assumption may not always hold true, potentially leading to residual confounding.In contrast, PSM creates a “quasi-experimental” setting by matching participants from the observation and control groups based on their propensity scores, which are derived from observed covariates. By ensuring that the distribution of multiple covariates is as similar as possible between the two groups, PSM better simulates the conditions of a randomized controlled trial. This approach effectively reduces selection bias and other systematic errors, thereby enhancing the robustness and reliability of the study conclusions [13, 14].The use of PSM in this study allowed for a more precise evaluation of each variable’s effect on IGRA false-negative results without necessitating specific assumptions about the relationships among covariates. This approach is especially important when investigating the complex interplay between biomarkers and clinical outcomes in intricate disease conditions [15]. By effectively balancing fundamental characteristics like age, gender, BMI, and other factors that might affect immune responses between the two groups, PSM ensured that any observed differences were more likely attributable to the variables of interest rather than confounding factors. This balanced design enhances the validity and reliability of the subsequent analyses, providing robust insights into the study’s objectives.

Following propensity score matching, this study observed that the observation group had significantly higher levels of WBC, NEUT, and NLR compared to the control group, whereas LYM, RBC, and ALB levels were notably lower. These differences suggest potential disparities in physiological states, inflammatory responses, or nutritional statuses between the two groups. Specifically, the elevated levels of WBC, NEUT, and NLR may indicate a more pronounced inflammatory response or immune activation in the observation group. Despite this heightened immune activity, it appears that the response was insufficient to effectively trigger the release of γ-interferon, which could explain the occurrence of false-negative results in the IGRA test.These findings highlight the complex interplay between immune activation and biomarker levels in patients with IGRA false negativity.

The inability to mount an effective γ-interferon response despite elevated inflammatory markers underscores the need for further investigation into the underlying mechanisms contributing to these discrepancies.Studies have found that excessive inflammatory responses may suppress specific immune response mechanisms, thereby affecting the outcomes of diagnostic tests [16]. Lower levels of LYM, RBC, and ALB could indicate impaired immune system function or malnutrition, which may weaken the body’s ability to combat tuberculosis infection.While clinical characteristics such as smoking history, alcohol consumption history, diabetes, hypertension, chronic lung disease, and pulmonary cavitation did not exhibit significant differences between the two groups, this does not diminish their importance. For example, diabetes has been shown to significantly alter host immune responses, making it more difficult to control Mycobacterium tuberculosis growth [17, 18]. The lack of significant differences in this study may be due to the specific sample characteristics or conditions under which the study was conducted. These factors might not have been the primary determinants of the observed differences between the groups within this particular study context.It is crucial to interpret these findings with caution. The absence of significant differences does not necessarily indicate a lack of impact. Factors such as the specific research background, sample size, and statistical methods employed can influence the results. Therefore, further investigation and consideration of these elements are warranted to fully understand the role of these clinical features in the context of IGRA false negativity.

Through multivariate binary logistic regression analysis, RBC and ALB were identified as influencing factors for IGRA false negativity, suggesting that decreased levels of RBC and ALB may be closely associated with the occurrence of false-negative IGRA results. Anemia and hypoalbuminemia, common manifestations of nutritional deficiencies, significantly reduce the body’s immune function, making patients more susceptible to pathogen invasion [19]. Malnutrition can also lead to reduced numbers and activity of immune cells, further compromising their ability to combat pathogens.This study’s findings are in line with those of previous research conducted by Wang MS et al. [20] and Sharninghausen JC et al. [21], who both reported that anemia and hypoalbuminemia are significant risk factors for false-negative IGRA results in tuberculosis patients. The consistency across these studies highlights the importance of considering nutritional status, such as anemia and low protein levels, when interpreting IGRA test results. These conditions can significantly impair immune function, leading to reduced sensitivity of the IGRA test and potentially affecting clinical decision-making.Based on these findings, this study constructed a nomogram model to predict false-negative IGRA results in PTB patients. The model demonstrated excellent goodness-of-fit, with an AUC of 0.703 from the ROC analysis, indicating robust predictive performance.

The model exhibited high accuracy, sensitivity, specificity, PPV, and NPV, further validating its reliability and practical utility.Decision curve analysis revealed that the net benefit of the nomogram model for predicting IGRA false negativity remained greater than zero when the threshold probability ranged from 0.15 to 0.75. This suggests that the model holds significant clinical application value, aiding in the improvement of PTB patient management strategies. By facilitating earlier diagnosis and treatment, the model can contribute to better public health outcomes.

### Limitations

This study has certain limitations. As a retrospective study, there may be unidentifiable or uncontrolled confounding factors that affect the generalizability and accuracy of the results. The relatively limited sample size and single-center origin may restrict the applicability of the study conclusions to broader populations. Additionally, while RBC and ALB were identified as influencing factors for IGRA false negativity, the causal relationship between these factors and IGRA test results remains unclear and requires further prospective studies for validation. This study primarily focused on hematological and biochemical indicators, future research should consider incorporating a wider range of biomarkers and clinical characteristics to comprehensively evaluate their impact on IGRA outcomes.

## Conclusion

In conclusion, this study employed Propensity Score Matching (PSM) to balance potential confounding factors between groups, thereby exploring the factors influencing false-negative IGRA results in pulmonary tuberculosis (PTB) patients. We successfully constructed a nomogram model with robust predictive performance, which can aid in optimizing early diagnosis and treatment decisions for PTB. This work provides new insights and methodologies for future research, potentially enhancing diagnostic accuracy and patient management strategies.

## Data Availability

All relevant data are within the manuscript and its Supporting Information files.

No

